# vPro-MS enables identification of human-pathogenic viruses from patient samples by untargeted proteomics

**DOI:** 10.1101/2024.08.21.24312107

**Authors:** Marica Grossegesse, Fabian Horn, Andreas Kurth, Peter Lasch, Andreas Nitsche, Joerg Doellinger

**Author notes:** corresponding author(s): Joerg Doellinger.

## Abstract

Viral infections are commonly diagnosed by the detection of viral genome fragments or proteins using targeted methods such as PCR and immunoassays. In contrast, metagenomics enables the untargeted identification of viral genomes, expanding its applicability across a broader spectrum. In this study, we introduce proteomics as a complementary approach for the untargeted identification of human-pathogenic viruses from patient samples. The viral proteomics workflow (vPro-MS) is based on an *in-silico* derived peptide library covering the human virome in UniProtKB (331 viruses, 20,386 genomes, 121,977 peptides), which was especially designed for diagnostic purposes. A scoring algorithm (vProID score) was developed to assess the confidence of virus identification from proteomics data (https://github.com/RKI-ZBS/vPro-MS). In combination with high-throughput diaPASEF-based data acquisition, this workflow enables the analysis of up to 60 samples per day. The specificity was determined to be > 99,9 % in an analysis of 221 plasma, swab and cell culture samples covering 18 different viruses (e.g. SARS, MERS, EBOV, MPXV). The sensitivity of this approach for the detection of SARS-CoV-2 in nasopharyngeal swabs corresponds to a PCR cycle threshold of 27 with comparable quantitative accuracy to metagenomics. vPro-MS enables the integration of untargeted virus identification in large-scale proteomic studies of biofluids such as human plasma to detect previously undiscovered virus infections in patient specimens.

## Introduction

Viruses are ubiquitous in the environment. They are known to infect all domains of life, including bacteria, fungi, plants and animals. Only a small proportion of these viruses can infect humans and cause disease. Increased research activity of viral infections as a result of the SARS-CoV-2 pandemic has once again emphasized, that an infection can have long-term consequences for patients and the society beyond the acute infection. Despite its importance, many viral infections are not diagnosed in detail. One of the consequences is that data on the surveillance of viral infections is incomplete and that long-term effects of past infections are difficult to recognize. Therefore, there is still a great need to develop diagnostic methods that provide a more comprehensive insight of viral infections in humans.

In general, methods for virus detection can be divided into two main categories, targeted and untargeted approaches. Targeted methods rely on known genetic markers, known proteins or other characteristics that are unique to a certain taxonomic level. They are suited for situations when an initial hypothesis about the virus causing the patient’s symptoms has been formulated or if a certain infection should be excluded. Currently, virus diagnostics is mostly based on targeted approaches, of which real-time *quantitative PCR* (qPCR) is the gold standard ^1, 2^. qPCR is characterized by high sensitivity, high specificity and ease of scalability and assay development. On-site diagnostics is dominated by immunoassays, such as lateral flow assays (LFA), which are cheap and rapid but usually less sensitive and potentially less specific than qPCR ^3, 4^. Both methods offer limited multiplexing capability. In contrast, untargeted approaches try to detect species without any prior knowledge or hypothesis regarding their identity. Untargeted identification of viruses can be achieved by metagenomic next-generation sequencing (mNGS), where viral sequences are identified within the total DNA/RNA content of a sample ^5, 6^. Viral metagenomics has paved the way towards a deeper understanding of the actual diversity of viruses in various environments and is used to diagnose infections with unknown etiology. Although, mNGS is gaining popularity, it is still limited by its sensitivity, its cost and the complexity of the laboratory and data analysis workflows ^7, 8^. In order to overcome these limitations NGS is often coupled with viral enrichment methods, e.g. PCR or hybridization probe capturing ^9-12^. Amplicon- and capture-based sequencing lacks the untargeted nature of mNGS but the multiplexing capacity is much higher as of conventional targeted methods. These methods play an important role in molecular epidemiology of viral infections as they allow to directly obtain whole virus genomes from patient samples while providing a higher sensitivity than mNGS ^7, 13^.

In microbiology, matrix-assisted laser desorption/ionization (MALDI) time-of-flight (ToF) mass spectrometry (MS) has become the method of choice for rapid, high-throughput and untargeted taxonomic classification of cultured bacteria. In virology, no comparable method exists, which would allow for fast, simple and inexpensive detection of viruses in an undirected manner. Protein analysis could theoretically be an alternative to NGS for untargeted virus detection. Due to technical limitations, such as low throughput, low sensitivity and lack of robustness, proteomics has not been widely used for this purpose. With the onset of the SARS-CoV-2 pandemic, an increasing number of studies were published in which targeted assays were developed, which enabled single virus detection by liquid chromatography and mass spectrometry (LC-MS) ^14-16^. Although great technical progress has been made in this direction, the technology is still inferior to current methods for targeted virus detection, especially qPCR, in terms of sensitivity and throughput ^17, 18^. Instead, we are convinced that the full potential of mass spectrometry for clinical diagnostics of viral infections is unfolded by the untargeted approach. Proteomics has made tremendous progress towards deep and comprehensive analysis of proteins at large-scale. The combination of data-independent acquisition (DIA), fast gradient liquid chromatography and AI-supported data analysis on the latest generation of MS instruments allows to identify thousands of proteins even in just 30 s ^19^. This technical progress has not been exploited for virus detection yet. Only very few proteomic studies exist, which aim to detect viruses in an untargeted or at least multiplexed approach ^20^. These studies rely on the use of data-dependent acquisition (DDA) on comparatively slow-scanning orbitrap instruments, which is currently being rapidly replaced for mere identification and quantification of proteins by DIA often performed on fast-scanning time-of flight (ToF) mass spectrometers. The high throughput and high sensitivity as well as the unbiased nature of precursor sampling makes DIA well suited for virus diagnostics.

In this study, a rapid proteomic workflow was developed, which enables the untargeted detection of human pathogenic viruses using DIA-MS and is named “vPro-MS” (**V**iral **Pro**tein Identification by **M**ass **S**pectrometry). All protein sequences of human pathogenic viruses (> 1.4 mio) in the UniProtKB were processed to construct a peptide spectral library (∼ 120,000 peptides, “vPro-MS spectral library”), which covers the entire human virome and is hence specifically designed for diagnostics. This library and its associated metadata were used to identify viral peptides in DIA-MS data and to subsequently assign the corresponding viruses, including highly-pathogenic species like monkeypox virus (MPXV) and Ebola virus (EBOV). The reliability of the virus identification is assessed by a score (“vProID score”) specifically adapted to untargeted proteomics. This proposed method can serve as a groundwork for virus diagnostics based on the latest technological developments in proteomics. In addition, the data analysis strategy can easily be adopted to detect virus-infected individuals in large-scale proteome cohort studies, thus contributing to the future understanding and monitoring of viral infections in the human population.

## Methods

### Samples

All samples containing viruses were handled in laboratories of biosafety level 2, 3 or 4 in accordance with their risk group classification. For specificity analysis, the following virus samples were used. Sucrose-cushion purified virus particles from cell-culture supernatant of Severe acute respiratory syndrome coronavirus type 2 (SARS-CoV-2, strain hCoV-19/Italy/INMI1-isl/2020, National Institute for Infectious Diseases, Rome, Italy, GISAID Accession EPI_ISL_410545), Vaccinia Virus (VACV, strain Western Reserve, GenBank GCA_900236015.1) and Cowpox Virus (CPXV, strain Brighton Red, GenBank GCA_000839185.1). Virus obtained from cell culture supernatant: Severe acute respiratory syndrome coronavirus (SARS-CoV, strain Hong Kong), SARS-CoV-2 (hCoV-19/Italy/INMI1-isl/2020, National Institute for Infectious Diseases, Rome, Italy, GISAID Accession EPI_ISL_410545), Middle East Respiratory Syndrome Coronavirus (MERS-CoV, kindly provided by Ron Fouchier, Erasmus University Rotterdam, Netherlands), Dengue Virus (DENV, serotype 1 ATCC VR-1856, serotype 2 ATCC VR-1584, serotype 3 ATCC VR-1256, serotype 4 ATCC VR-1490), Human Coronavirus OC43 (HCoV-OC43,ATCC CCL-244), Human Coronavirus 229E (HCoV-229E ATCC CCL-171), Human Coronavirus NL-63 (HCoV-NL63, BEI Resources, NIAID, NIH: Human Coronavirus, NL63, NR-470), Monkeypox Virus (MPXV,WHO/INSTAND 7418A-230516-01 to −05). Lassa Virus Hawa Forray (LASV, Rocky Mountain Laboratories, NIH, USA), Reston Ebolavirus USA (RESTV, Philipps University Marburg, Germany), Zaire Ebolavirus Mayinga (EBOV, Philipps University Marburg, Germany), Marburg Virus Musoke (MARV, Philipps University Marburg, Germany).

For sensitivity analysis, a well-established panel consisting of 66 SARS-CoV-2 swab samples (with different Ct values was used ^21^. This panel had been previously used for validating SARS-CoV-2 antigen rapid tests in a larger scale in different laboratories in Germany. SDS (20 % solution, Calbiochem) was added to each virus-containing sample to a final concentration of 1 %. Subsequently, the samples were either heated to 100°C for 5 min (LASV, RESTV, ZEBOV, MARV) or heated to 95°C for 10 min (other viruses) and stored at −20 °C until further use.

### Samples Preparation for Proteomics

All samples (swab samples, cell culture supernatants, purified viruses) were provided as liquids. Reduction and alkylation of cysteines was not included due to time constraints in this study but could potentially be integrated at this point as the peptide library provided with this study contains carbamidomethylated cysteines. Proteins were processed using S-Trap™ micro columns (Protifi, Fairport, NY) according to the manufacturer’s instructions ^22^. Proteins were digested for 1 h at 47 °C using 2 μg Trypsin Gold, Mass Spectrometry Grade (Promega, Fitchburg, WI) per sample in 50 mM TEAB. After elution, peptides were evaporated to dryness, resuspended in 20 μL 0.1% formic acid and quantified by measuring the absorbance at 280 nm using the NanoPhotometer® NP80 (Implen, Munich, Germany).

### Liquid chromatography and mass spectrometry

Peptides were analyzed on an Evosep One liquid chromatography system (Evosep, Odense, Denmark) coupled online via the CaptiveSpray source to a timsTOF HT mass spectrometer (Bruker Daltonics, Bremen, Germany). 1 μg of peptides was manually loaded onto Evotips Pure (Evosep) and separated using the 60 samples per day (SPD) method on the respective performance column (8 cm x 150 μm, 1.9 μm, Evosep). Column temperature was kept at 40 °C using a column toaster (Bruker Daltonics) and peptides were ionized using electrospray with a CaptiveSpray emitter (20 μm i.d., Bruker Daltonics) at a capillary voltage of 1750 V. Spectra were acquired in diaPASEF mode in the m/z range of 100-1,700 and in the ion mobility (IM) range of 0.65 – 1.35 Vs/cm^2^. Each scan cycle was comprised of 12 diaPASEF scans, which consisted of two IM windows with variable isolation window widths adjusted to the precursor densities using py_diAID covering the m/z range of 350 – 1,150 ^23^. The collision energy was decreased as a function of the IM from 59 eV at 1/K0 = 1.6 Vs/cm to 20 eV at 1/K0 = 0.6 Vs/cm. The accumulation and ramp times were specified as 100 ms.

### vPro-MS Peptide Spectral Library

Protein sequences of all human pathogenic viruses (search term: “(taxonomy_id:10239) AND (virus_host_id:9606)”) were downloaded from UniProt (release 2023_01, 1,463,727 entries) ^24^. Gene ontology and taxonomic information of the proteins were also retrieved from Uniprot. At first, the protein sequences were filtered to keep only structural viral proteins according to their gene ontology (GO) annotation (capsid, envelope, virus membrane), which were part of a complete viral proteome using R (version 4.1.3). Afterwards, a peptide library of the remaining protein sequences was predicted using the deep-learning algorithm implemented in DIA-NN (version 1.8.1) with strict trypsin specificity (KR not P) allowing no missed cleavage site in the m/z range of 350 – 1,150 with charges states of 2 – 4 for all peptides consisting of 7-30 amino acids with enabled N-terminal methionine excision and cysteine carbamidomethylation ^25^. The resulting peptides in the spectral library were filtered in R (version 4.1.3) based on retention time (iRT: −40 – 130) and ion mobility (1/K_0_: 0.65 – 1.35 Vs/cm^2^). The remaining peptides were taxonomically annotated based on the organism information of UniProt, which was manually curated to enhance data integrity. Afterwards, the following peptides were removed: (i) peptides, which were not specific to a certain virus species, (ii) peptides mapping to the human proteome, (iii) peptides mapping to common protein contaminants (BSA, trypsin, keratins). Information on the remaining sequences (121,977) was summarized into three separate output files. The “Taxonomy.Summary” file summarizes the taxonomic information of the library and contains data about the number of proteomes per virus species as well as the median and maximum number of unique peptides per proteome and the sum of all unique peptides per species. The “vPro.Peptide.Library” file contains data for each peptide, including spectral library information (rt, IM, mz, modifications), taxonomic information (species, subspecies) and protein-specific information (gene name, protein Name, UniProtID). It served as the database for virus identification. Furthermore, a “vPro.Virus.fasta” file was created, in which each peptide sequence has its own entry with a unique identifier. This file was merged with a human (UniProt, UP000005640) and a contaminant peptide fasta file, which was used for predicting the final spectral library by using the deep-learning algorithm implemented in DIA-NN (version 1.8.1) ^25^. In order to predict a spectral library from the peptide sequence file, the additional commands “--cut” and “--duplicate-proteins” were used.

### Peptide Identification

The LC-IMS-MS/MS data were analyzed in DIA-NN using default settings including a false discovery rate (FDR) of 1 % for precursor identifications with disabled “match between run” (MBR) option to receive independent outputs for each sample. Mass calibration was disabled and mass tolerance was set to 15 ppm for MS^1^ and MS^2^ spectra. The resulting main report was used for virus detection.

### vPro-MS – Virus Identification

The vPro-MS virus identification script was written in R (version 4.1.3) and can be downloaded from [github] (https://github.com/RKI-ZBS/vPro-MS). At first the main report from DIA-NN is loaded and precursor information is collapsed into peptides after applying a filter (minimum Cscore of 0.95). Viral peptides are identified and annotated with taxonomic, proteome- and protein-related information using the “vPro.Viral.Peptide.Library” file. A virus identification score (vProID score) is then calculated according to the following equation:

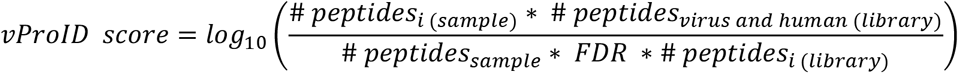

i = virus proteomes in library

The vProID score is the log10 value of the number of identified peptides for a given virus proteome in a certain sample divided by the expected number of peptide identifications for this virus proteome just by chance in this specific sample. vProID score is calculated independently for each virus proteome in the human virome library with a matching peptide sequence in the DIA-NN results. The threshold for virus identification used in this study was set to a minimum of 2 peptides and a vProID score of 2. A vProID score of 2 means, that the number of identified peptides for a given viral proteome exceeds the number expected by chance 100-fold. Afterwards information for all species with at least one proteome above the threshold is aggregated into an output table reporting the viral species and subspecies identified in each sample in conjunction with all matched peptide sequences and the top-ranked proteome. Viruses are quantified using a Top3 approach ^26^.

### Synthetic SARS-CoV-2 Peptide Spectral Library

All peptides specific for SARS-CoV-2 in the vPro-MS spectral library (n= 75) were synthesized by JPT (PT Peptide Technologies, Berlin, Germany) either as SpikeTide or MaxiSpikeTide peptides. Afterwards 100 fmol of each peptide was spiked into 8 different nasopharyngeal swab samples obtained from healthy individuals, which were analyzed by LC-IMS-MS/MS using the same settings as described above. The resulting data were analyzed in DIA-NN. At first, a spectral library was predicted from human and SARS-CoV-2 peptides of the vPro-MS library using DIA-NN’s deep-learning algorithm with the additional commands “--cut” and “--duplicate-proteins”. Precursors were identified from the 8 samples in a combined analysis using the predicted library with default settings and exported as a spectral library, which contained 68 SARS-CoV-2 peptide sequences. This spectral library was used to re-analyze the data from the SARS-CoV-2 panel using default settings in DIA-NN.

## Results

### vPro-MS workflow for rapid and untargeted virus identification

The vPro-MS workflow for untargeted virus detection by proteomics consists of three main parts: sample preparation, LC-MS measurements and data analysis (Fig. 1). The workflow takes about 2h to perform from sample preparation to result (vPro-MS report) and has a throughput of 60 samples per day (SPD). Samples analyzed in this study were prepared using a slightly modified STrap protocol and the resulting peptides were loaded manually onto EvoTips. The 1 h digestion of proteins into peptides is the longest step in the sample preparation procedure and the whole workflow. In general, alternative sample preparation strategies should work similarly well, but care must be taken to select lysis buffers and conditions, which effectively inactivate viruses ^27^. Samples were analyzed using the 60 SPD method on an Evosep LC system (24 min per sample) and a diaPASEF data acquisition scheme on a timsTOF HT mass spectrometer. This LC-MS system offers the robustness necessary to measure thousands of samples in a routine environment. Peptide sequences were identified from the LC-MS data using DiaNN and the vPro-MS spectral library of the human virome. Human-pathogenic viruses were detected from the DiaNN output using the vPro-MS R script and meta data of the vPro-MS spectral library. The reliability of virus detection is monitored by calculating a confidence score (vProID) and the results are summarized in a tabular report. Samples can be analyzed in a batch mode in which the peptide identification with DIA-NN is the speed-limiting step. This step can be computationally parallelized. In our computational setting (Intel® Xeon® Platinum 8160, 24 cores, 192 GB RAM) the data analysis was performed regularly in less than 10 minutes per sample.

**Figure 1:**
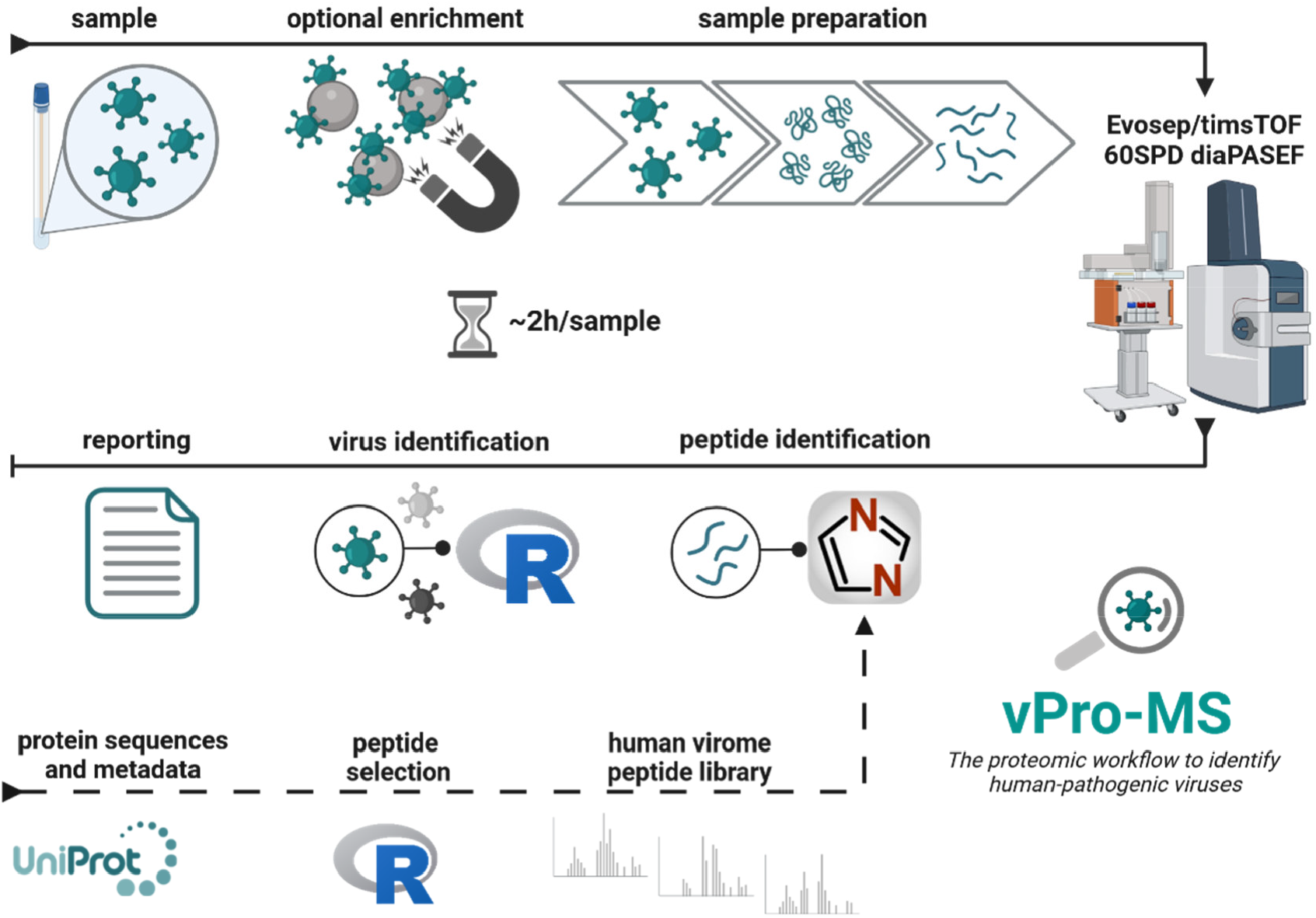
Overview of the vPro-MS workflow for virus identification by untargeted proteomics. The vPro-MS workflow for virus detection by untargeted proteomics begins with the sample preparation. Proteins are digested into tryptic peptides using S-Traps and loaded onto Evotips. Afterwards peptides are analyzed for 24 min per sample (corresponding to a throughput of 60 samples per day) using diaPASEF on an Evosep One coupled to a timsTOF HT mass spectrometer. Peptide sequences are identified from the MS data using DIA-NN (v 1.8.1) with the vPro-MS peptide library, which is based on UniProt. These peptide sequences are further analyzed by the vPro-MS R script to identify human-pathogenic viruses and generate the vPro-MS report. The confidence of virus identification is assessed by the vProID score. *(Created in BioRender. Grossegesse, M. (2024) BioRender.com/s15u220)*

### Construction of the human virome peptide spectral library

The strategy of the vPro-MS data analysis workflow is to construct a peptide spectral library covering the complete human virome (Fig. 2). This library is then used to identify peptide sequences in DIA-MS data. The peptide spectral library was constructed from all protein sequences of human-pathogenic viruses available in the UniProt Knowledgebase (1,463,727 proteins, release: 2023_1). Initially, proteins without an associated proteome ID were removed. Structural proteins are best suited for the detection of viruses, as they are the most abundant of all virus proteins. Therefore, proteins were filtered according to their GOCC (Gene Ontology Cellular Component) terms and only structural proteins, such as core, envelope and virus membrane proteins were retained (49,657 proteins). An *in-silico* peptide library was predicted from those proteins using DIA-NN to filter the resulting tryptic peptides according to their detectability in terms of m/z values, retention time and ion mobility (126,788 peptides). The lowest common ancestors (lca) of those peptides were analyzed in R using taxonomic information from UniProt and only species-specific peptides were kept. Taxonomic information below the species rank was aggregated into a subspecies rank. Peptides matching to either the human proteome or common contaminants, such as BSA or trypsin, were removed and the remaining sequences were annotated with protein-level (proteomeID, UniprotID, gene name, protein name) precursor-level (m/z, charge state, iRT, IM, M(ox), CAM) and taxonomic (species, subspecies) information. The peptides were exported as a table (Viral.Peptide.Library.txt), which served as the database for virus detection in DIA-NN precursor output tables. The library consisted of 121,977 peptide sequences from 331 human-pathogenic viruses and was constructed from 20,386 virus proteomes. The peptides were also exported in .fasta format (VirusID.fasta) along with peptide .fasta files of the human proteome and common contaminants. These files were used to predict a peptide spectral library of the human virome in DIA-NN (Library_VirusID_v3-lib.predicted.speclib). A detailed taxonomic summary of the library is provided in Table S1.

**Figure 2:**
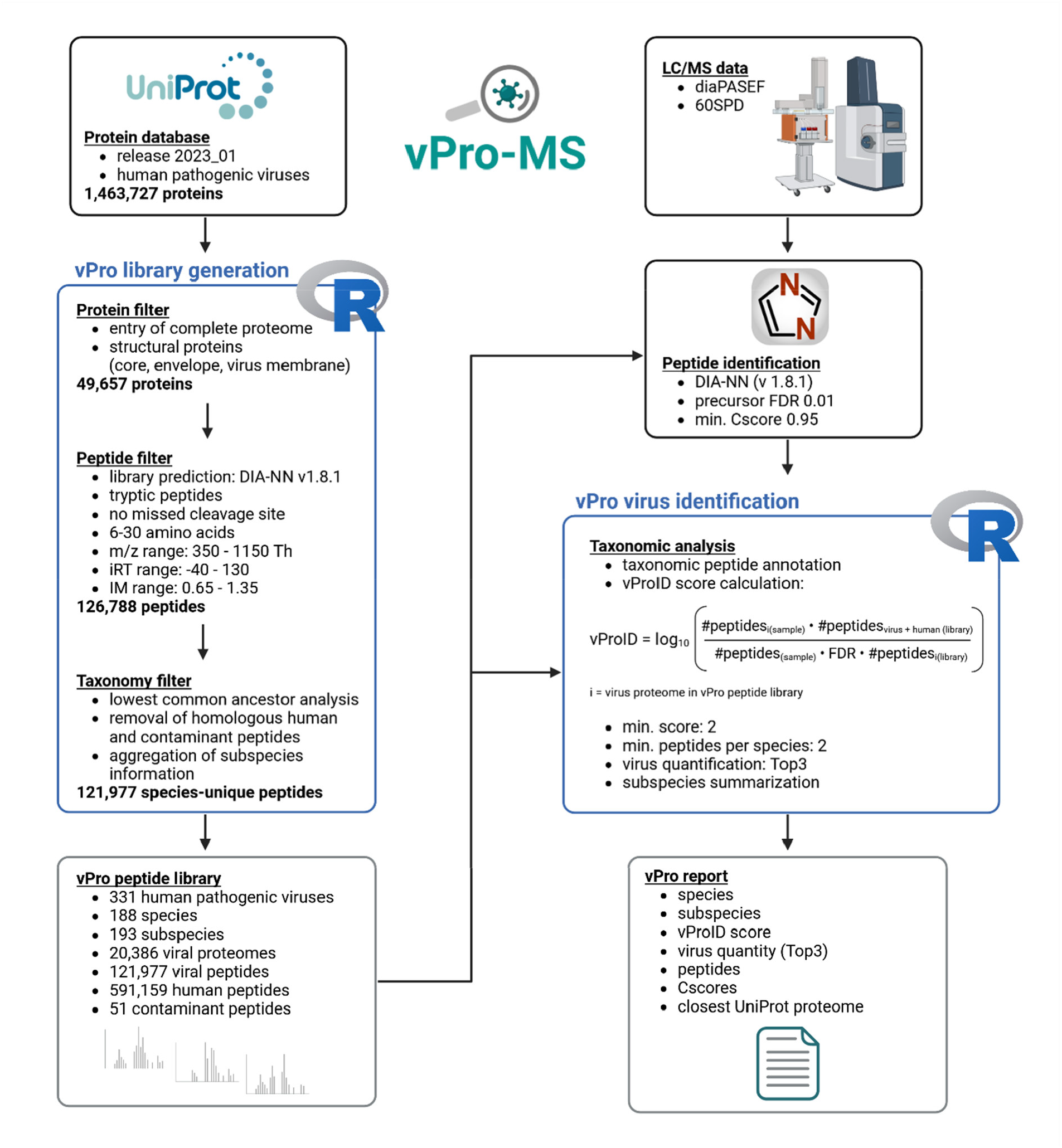
Data flow chart of vPro-MS for virus detection by proteomics. Data processing of the vPro-MS workflow splits into two parts, peptide library construction and virus detection. At first, a peptide library is generated based on UniProt protein sequences. The UniProt database (release 2023_01) contains > 1.4 million protein sequences from human-pathogenic viruses. Structural proteins are extracted from these sequences and are used to predict a peptide spectral library. Peptides are further filtered for detectability (m/z, iRT, IM) and taxonomic specificity. The remaining peptides form the vPro-MS peptide library, to which human and contaminant sequences are added. This library is used to identify peptides from DIA-MS data using DIA-NN. The peptide sequences are analyzed using the vPro-MS R script to identify human-pathogenic viruses. vPro-MS controls the reliability of virus detection is by calculating a confidence score (vProID) and summarizes the results in a tabular report. *(Created in BioRender. Grossegesse, M. (2024) BioRender.com/x53q677)*

### Assessing the confidence of virus identification

The vPro-MS peptide spectral library covering the complete human virome was used to identify peptide sequences in DIA-MS data. This strategy posed a major challenge for post-processing of the identification results in order to achieve a high specificity when trying to identify very few viral peptides in a sample within a library of more than 100,000 viral sequences. Therefore, confidence of virus identification was supervised with the development of the vProID score.

The number of specific peptides varied greatly between the different species within the library. HIV-1 was by far the largest part of the library with 68,103 peptides which equals 56 % of all entries. As peptides are usually identified in proteomics with a controlled error rate (often ∼ 1 %), this means that few HIV-1 sequences were identified in almost every sample just by chance. In order to control the reliability of virus identification, we introduced a confidence score, named vProID (**V**iral **Pro**tein **Id**entification). Care was taken to ensure that the sensitivity of virus detection was not impaired. This score is the log10 value of the number of identified peptides for a certain virus proteome divided by the number of expected peptides for this virus proteome just by chance in a specific sample. Afterwards the top ranked proteome for each virus species was kept and a vProID threshold was applied to filter out unreliable virus identifications. Proteomes with a single matching peptide were also removed at this step. An example for filtering the top ranked virus proteomes per species based on the vProID score is provided in table S2. The discriminatory power of the vProID score to distinguish between random and true virus identifications for the analysis of nasal swab samples is shown in Fig. 3. In total, 58 swab samples positive for SARS-CoV-2 covering a ct-range of 18 - 35, and 8 SARS-CoV-2 negative swab samples were tested for 331 human-pathogenic viruses, which is equivalent to 21,846 PCR tests. Specificity on virus level was used to determine the outcome and was defined as the number of true negative (TN) virus tests divided by the sum of true negative and false positive (FP) virus tests (specificity_Virus_ = TN/(TN+FP). When applying a 1 % FDR on precursor-level, 959 viruses were incorrectly identified, which equals ∼ 14 viruses per sample. An increasing stringency of the cut-offs for precursor identifications led to the reduction of incorrect virus identifications, but only the combination of 0.1 % FDR and at least 2 peptide sequences per virus was able to achieve a specificity of 100 %. However, this increase in specificity corresponds to a loss of sensitivity. The vPro-MS data analysis using the vProID score also achieves a specificity of 100 % but has a higher sensitivity for the detection of SARS-CoV-2 than using the most stringent identification threshold (ct 27 vs 26.5). These results underline, that the vProID score is able to separate random from true virus identifications even when using a rather moderate FDR of 1 % for a dataset of 807,221 (redundant) peptide identifications.

**Figure 3:**
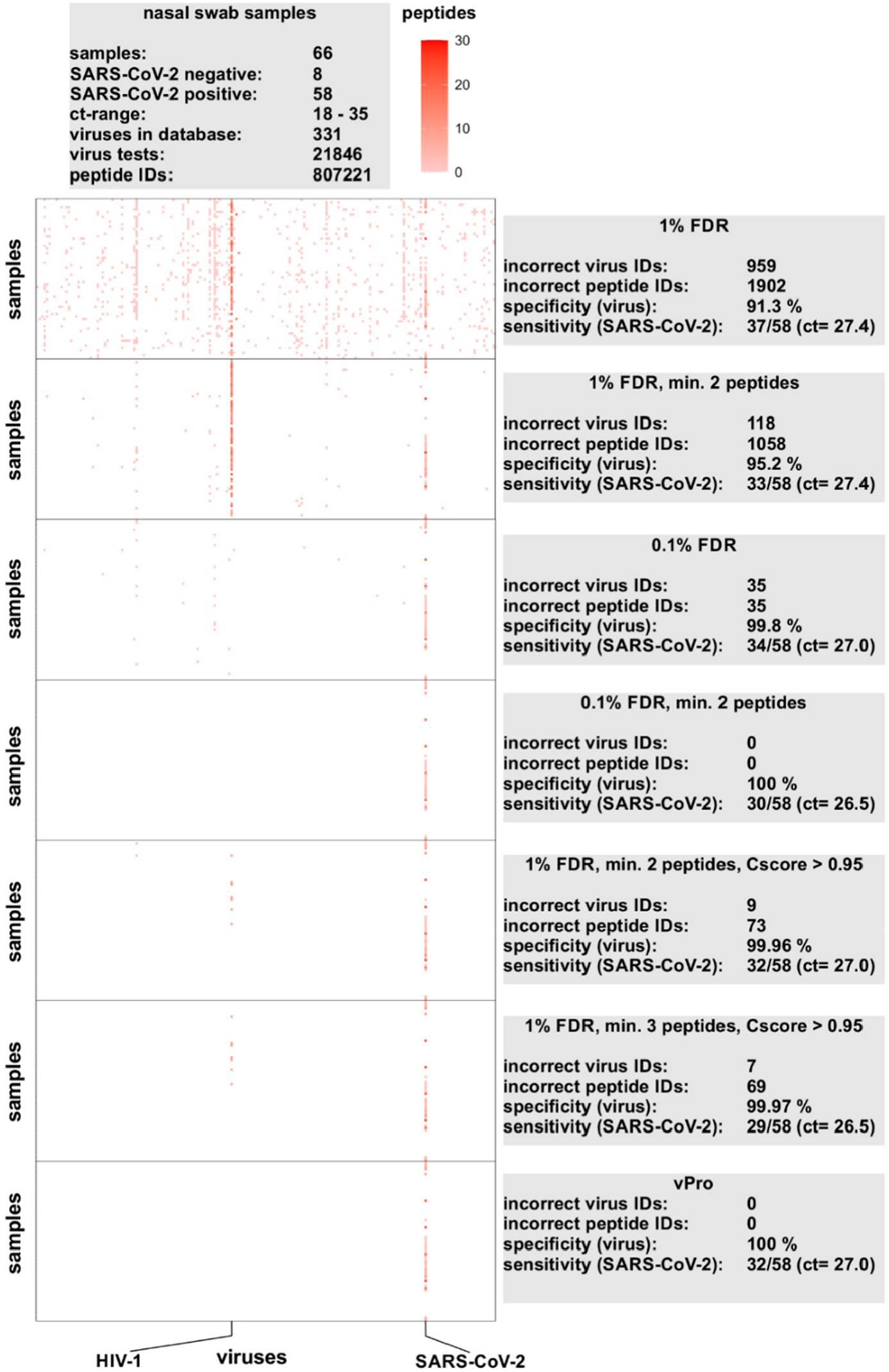
Comparison of different identification thresholds for virus detection. The vPro-MS peptide spectral library covering the human virome (331 viruses) was used to identify peptides in 66 nasal swab samples, of which 58 were positive for SARS-CoV-2 (ct range 18 - 35) and 8 were negative. This corresponds to 21,846 individual virus tests within a dataset consisting of 807,221 (redundant) peptide identifications. Afterwards, different virus identification thresholds were applied to the DIA-NN output. The heatmap visualizes the number of identified peptides for each of the 331 viruses in alphabetical order (x-axis) and samples (y-axis). The results for different filter parameters are summarized in the grey boxes, including the number of incorrect virus and peptide identifications as well as the specificity calculated on virus level. The sensitivity is shown as the number of SARS-CoV-2 positive identifications by vPro-MS in relation to the number of SARS-CoV-2 positive samples defined by qPCR as well as the respective PCR cycle threshold (ct) (95% CI).

### Evaluation of the specificity of vPro-MS for virus detection

The specificity of the vPro-MS workflow was determined by the analysis of 221 samples from 4 different sources covering 18 human-pathogenic viruses (Fig. 4). Two sample panels were analyzed for this study and the raw data from two further studies of patient samples were downloaded from the PRIDE repository ^28, 29^. The sample types included purified viruses, cell culture supernatants, nasal swabs and plasma (Fig. 4A). MS data of all samples were analyzed using the vPro-MS data workflow covering 331 human-pathogenic viruses. This corresponded to the analysis of 73,151 individual virus tests. Specificity was calculated either on virus or sample level as the number of true negative (TN) results divided by the sum of true negative and false positive (FP) results (specificity = TN/(TN+FP). The specificities ranged from 99,97 – 100 % on virus level and from 95,74 – 100 % on sample level (Fig. 4B). The analysis of the specificity panel revealed 3 additional virus species identifications in one monkeypox and one vaccinia virus sample (Fig. 4C). These two viruses belong to the same virus family of poxviruses, genus orthopoxviruses. In both cases, the top hit based on the virID confidence scores was the correct orthopoxvirus species. However, in both samples either one or two closely related orthopoxvirus species were identified as well. Most probably, this resulted from the incompleteness of the sequence database underlying the vPro-MS peptide library. It might occur, that certain tryptic peptides were considered to be species-unique but in fact the respective sequence actually occurs also in closely-related species, for which the respective isolates are missing in the database. Therefore, care should be taken if several species of the same genera are identified in one sample. In the nasal swab study of SARS-CoV-2 patients obtained from PRIDE, SARS-CoV-2 was detected in two samples, which were labeled negative. However, as this study was about analyzing SARS-CoV-2 positive samples, it is unlikely, that these additional identifications were random events. If this would be true, it must have been expected that any other species would have been identified. Therefore, it is more likely, that those additional identifications were either due to peptide carry-over between samples or were resulting from an error in the initial characterization of those samples. Overall, the data underlined that the vPro-MS data analysis workflow is highly specific for the detection of human-pathogenic viruses in different sample types and data from various laboratories.

**Figure 4:**
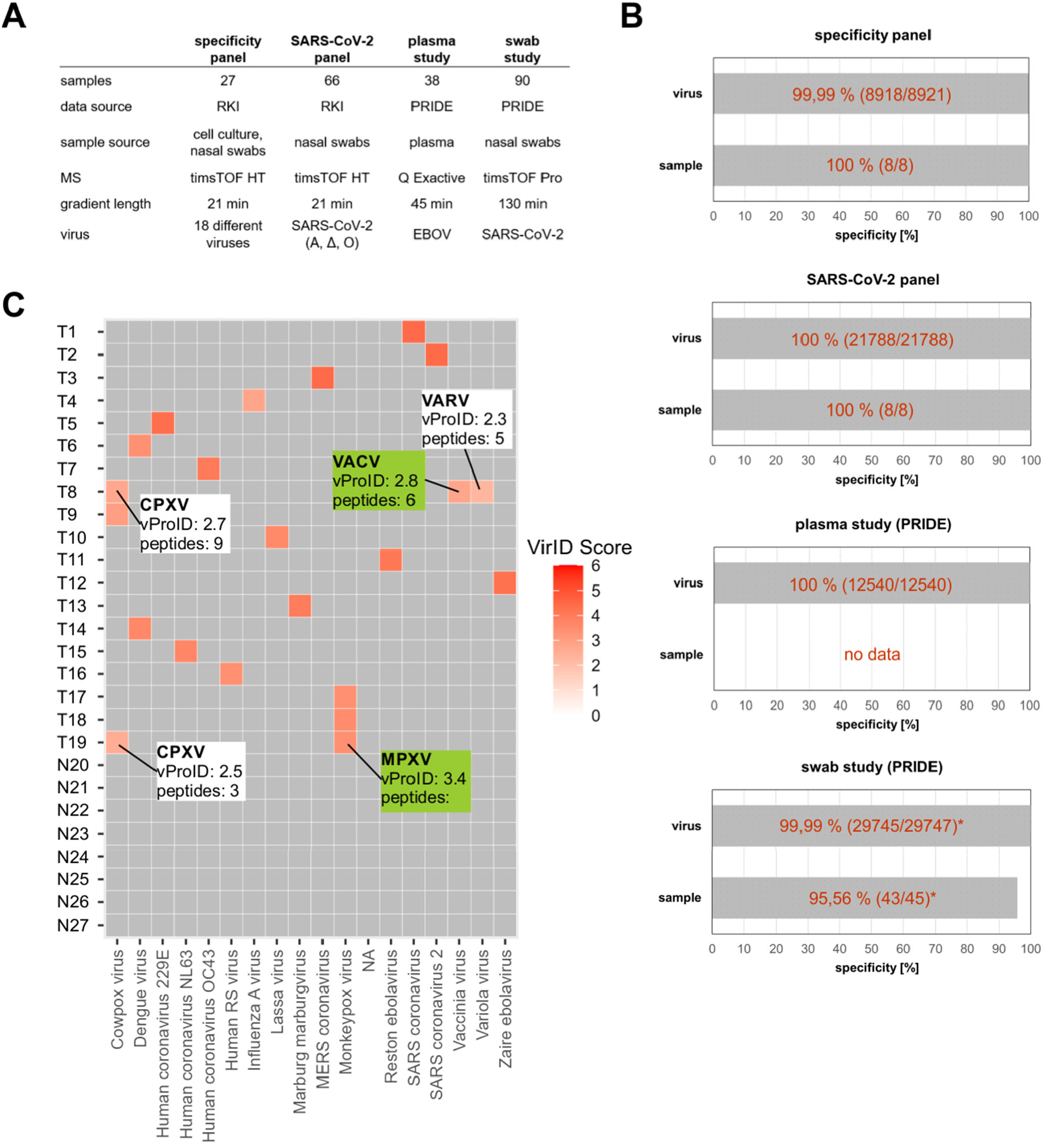
Evaluation of the specificity of vPro-MS for virus detection. The specificity of the vPro-MS workflow was evaluated by analyzing 221 samples from 4 sources covering 18 different human-pathogenic virus species (A). Two sample panels were analyzed by MS for this study and the raw data from two further studies of patient samples were downloaded from PRIDE^28, 29^. The sample types included cell culture supernatants, nasal swabs and plasma. MS data of all samples were analyzed using the vPro-MS data workflow covering 331 human-pathogenic viruses. This corresponded to the analysis of 73,151 individual virus tests. The specificities ranged from 99,97 – 100% on virus level and from 95,74 – 100 % on sample level. The number of correct classifications in relation to the total number of tests is given in brackets (B). The results of the specificity panel are further visualized in a heatmap, which displays the vProID score for each species and sample (C). For two orthopoxvirus samples (T8 and T19) with additional virus identifications of closely-related species, further information is provided in white and green boxes.

**Figure 5:**
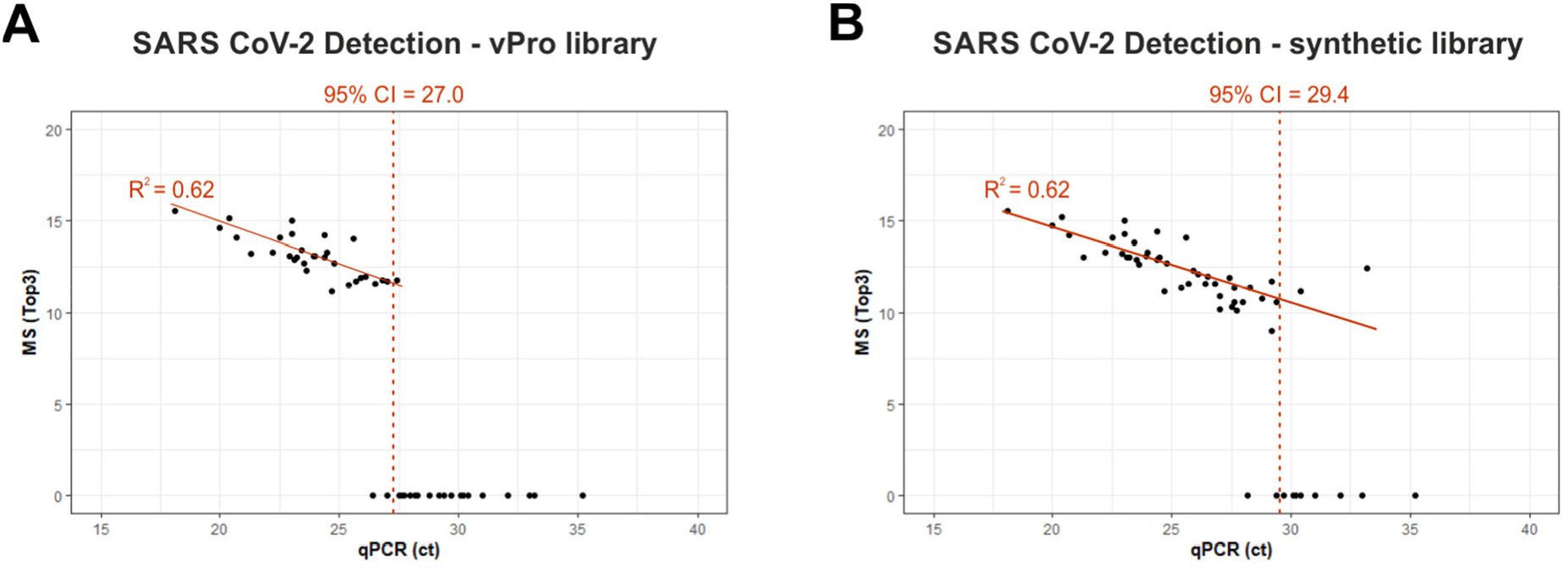
Evaluation of the sensitivity of vPro-MS for the detection of SARS-CoV-2 in nasal swabs. The sensitivity of vPro-MS for the detection of SARS-CoV-2 was evaluated in 66 nasal swab samples covering a Ct range of 18 - 35. The panel included 8 negative samples and 58 SARS-CoV-2 positive samples of three different variants (alpha, delta, omicron). The quantitative values for proteomics (Top3) and qPCR (ct) were compared when using the vPro-MS peptide library (A) and a library constructed from analysis of synthetic SARS-CoV-2 peptides (B), including all theoretical peptides of the SARS-CoV-2 nucleoprotein within the vPro-MS spectral library. The limits of detection are displayed at 95 % confidence.

### Evaluation of the sensitivity of vPro-MS for the detection of SARS-CoV-2

The sensitivity of the vPro-MS workflow for the detection of SARS-CoV-2 was evaluated in 66 nasal swab samples covering a Ct range of 18 - 35. The panel included 58 SARS-CoV-2 positive samples of three different variants (alpha, delta, omicron) and additionally 8 SARS-CoV-2 negative samples. The limit of detection (LOD) with 95 % confidence corresponded to a Ct value of 27 as determined by qPCR. It should be noted that approximately twice the amount of starting material (27 μL) was used per qPCR reaction compared to an average volume of 12 μL starting material, which was injected per proteomics measurement. However, twice the sample amount corresponds to a very small Ct difference of just 1 Ct. Virus quantification by proteomics and qPCR correlated with an R^2^ value of 0.62. In order to increase the sensitivity, we evaluated the use of a synthetic SARS-CoV-2 library for enhanced peptide identification. Therefore, all theoretical peptides of the SARS-CoV-2 nucleoprotein were synthesized and spiked into negative swab samples to create a SARS-CoV-2 peptide spectral library including human nasal swab peptides. This library improved the LOD with 95 % confidence to a Ct value of 29.4, which is equivalent to ∼ 5 times less virus being detected compared to the predicted library. This underlines the potential of library construction from synthetic peptides to improve the sensitivity in the future. Such specific libraries can be constructed from any sample, in which viruses were identified by the vPro-MS peptide library of the human virome.

## Discussion

The vPro-MS workflow enables untargeted proteomics to detect human-pathogenic viruses with high specificity in various sample types. Currently, untargeted virus detection is solely based on nucleic-acid analysis using mNGS. Protein analysis adds some complementary benefits to the virological toolbox. Proteins are more stable than RNA, which could enable virus detection in samples unsuitable for mNGS. The stability of proteins is the main reason for the increasing interest for proteomics of ancient proteins ^30^. The conservation of viral genome sequences can be rather low for certain species, which can complicate the detection of virus variants. This is the reason why amplicon and capture sequencing methods using NGS require frequent adaptations. Protein sequences are more stable than viral genomes as mutations are only partially translated into gene products, which might be beneficial for virus detection in certain cases. Furthermore, mNGS workflows often focus on either RNA or DNA, which are the molecular building blocks of viral genomes. As all viruses contain proteins, the analysis of DNA and RNA viruses with vPro does not require a customized laboratory and data analysis pipeline. Untargeted proteomics has therefore the capability to enable a high-throughput screening of the human virome with little prior knowledge or customizations.

We evaluated the performance of vPro-MS by collecting data of the sensitivity, specificity and quantitative correlation with qPCR from published mNGS experiments for the detection of SARS-CoV-2 in swab samples (Table S3) ^7, 31-34^. The comparison of mNGS and proteomics for the detection of SARS-CoV-2 in swab samples reveals, that proteomics is already able to provide a similar specificity on species-level and a comparable quantitative correlation to qPCR as mNGS at similar or even higher throughput. The proteomics workflow is characterized by its comparatively simple sample preparation with low reagent costs and fast and simple data analysis. The LC-MS system used in this study minimizes the risk of cross-contamination due to the disposable sample spin filters and disposable LC trap columns. However, it is essential to wash the LC columns between samples. Samples are measured and analyzed in a consecutive order. Therefore, the workflow omits the risk of cross-contamination introduced by multiplexing, which is common in mNGS, and provides a high flexibility as sample orders can be prioritized ^35^. While it is uneconomical to sequence few numbers of urgent samples on a high-throughput NGS platform, in proteomics cost and time scale linearly with sample numbers. An urgent sample can be analyzed in 2 h from swab to report using the vPro-MS workflow whenever needed. The throughput of proteomics is currently limited by the gradient length needed to achieve a certain analysis depth. However, the number of analyzed proteins per time is steadily increasing and has already reached ∼ 1,500 unique protein identifications per minute ^36^. This continuing progress will increase the throughput of proteomics-based virus diagnostics in the future. A further potential benefit of proteomic-based virus diagnostics compared to NGS-based analysis is that proteomics simultaneously provides information about the patient’s immune response, which is encoded in the host proteome and not in the host genome.

The current limitation for virus detection using untargeted proteomics is the sensitivity. The conclusions drawn from the comparison of sensitivities between different methods applied to different sample cohorts are very limited (Table S3). The sensitivities in the mNGS studies were reported as the proportion of samples correctly identified as positive. This does not account for differences in Ct distribution within the sample cohorts. Therefore, the median Ct values should be considered as well. Based on our overview, we hypothesize, that proteomics is in its current stage of development less sensitive than mNGS. However, it should be noted that the sensitivity of mNGS but also of proteomics depends on the selected throughput. The main focus to develop proteomics into a widely adopted technology for untargeted virus detection must therefore be the improvement of its sensitivity. The sensitivity of vPro-MS will directly benefit from recent technological improvements in proteomics, such as new MS instruments, as the workflow is based on DIA-MS, which has become the most popular and most promising analysis strategy in proteomics ^37^. In this study, we demonstrated the possibility of improving the sensitivity of our approach by optimizing the peptide spectral library. We were able to increase the sensitivity to detect SARS-CoV-2 in swab samples 5-fold by constructing a library from synthetic viral peptides. The determined LOD of Ct 29,4 is close to the expected threshold for infectivity of ∼ Ct 30 ^38, 39^. We expect that the overall sensitivity of our approach will be increased further if we can synthesize all remaining peptides from the human virome. This would require a significant reduction of the human virome library of vPro. This could be achieved by the use of sequence conservation metrics and of artificial intelligence-based prediction of peptide detectability. Another large and untapped potential lies in the adoption of depletion or enrichment strategies during sample preparation, which is common in mNGS ^40^. For proteomics such a strategy has not been developed. This is especially important as many virus samples are stored in virus transport medium (VTM) to preserve virus specimens after collection. This medium contains very large amounts of proteins, namely albumin and collagen, which severely reduce the sensitivity of proteomics. The situation is comparable to plasma proteomics, which has been struggling with a large dynamic range for many years. Recently, several sample preparation protocols were developed, which are able to overcome this limitation ^41, 42^. Similar strategies could be used to improve the sensitivity of proteomics for untargeted virus detection.

Taken together, this study introduces proteomics as an alternative approach for untargeted identification of human viruses. We show, that our proposed workflow is flexible, rapid and offers high specificity for virus detection in different sample types, such as plasma and swabs. vPro-MS has a high potential to further improve its sensitivity, which should enable its wider adoption for high-throughput and rather low-cost screening studies. The data analysis workflow of vPro-MS can further be integrated into large-scale proteome studies of biofluids such as human plasma to explain outliers due to acute infections and to determine the prevalence of persistent infections, such as SARS-CoV-2 ^42^.

## Data Availability

All data produced in the present study are available upon reasonable request to the authors

## Acknowledgements

The authors thank Andreas Puyskens, Caroline Eberle and Juliane Fraissinet for providing the viruses.

## References

(1) Cassedy, A.; Parle-McDermott, A.; O’Kennedy, R. Virus Detection: A Review of the Current and Emerging Molecular and Immunological Methods. Front Mol Biosci 2021, 8, 637559. DOI: 10.3389/fmolb.2021.637559 From NLM PubMed-not-MEDLINE.

(2) Dutta, D.; Naiyer, S.; Mansuri, S.; Soni, N.; Singh, V.; Bhat, K. H.; Singh, N.; Arora, G.; Mansuri, M. S. COVID-19 Diagnosis: A Comprehensive Review of the RT-qPCR Method for Detection of SARS-CoV-2. Diagnostics (Basel) 2022, 12 (6). DOI: 10.3390/diagnostics12061503 From NLM PubMed-not-MEDLINE.

(3) Scheiblauer, H.; Filomena, A.; Nitsche, A.; Puyskens, A.; Corman, V. M.; Drosten, C.; Zwirglmaier, K.; Lange, C.; Emmerich, P.; Muller, M.; et al. Comparative sensitivity evaluation for 122 CE-marked rapid diagnostic tests for SARS-CoV-2 antigen, Germany, September 2020 to April 2021. Euro Surveill 2021, 26 (44). DOI: 10.2807/1560-7917.ES.2021.26.44.2100441 From NLM Medline.

(4) Puyskens, A.; Bayram, F.; Sesver, A.; Michel, J.; Krause, E.; Bourquain, D.; Filomena, A.; Esser-Nobis, K.; Steffanowski, C.; Nubling, C. M.; et al. Performance of 20 rapid antigen detection tests to detect SARS-CoV-2 B.1.617.2 (Delta) and B.1.1.529 (Omicron) variants using a clinical specimen panel from January 2022, Berlin, Germany. Euro Surveill 2023, 28 (16). DOI: 10.2807/1560-7917.ES.2023.28.16.2200615 From NLM Medline.

(5) Gauthier, N. P. G.; Chorlton, S. D.; Krajden, M.; Manges, A. R. Agnostic Sequencing for Detection of Viral Pathogens. Clin Microbiol Rev 2023, 36 (1), e0011922. DOI: 10.1128/cmr.00119-22 From NLM Medline.

(6) Duan, H.; Li, X.; Mei, A.; Li, P.; Liu, Y.; Li, X.; Li, W.; Wang, C.; Xie, S. The diagnostic value of metagenomic next rectanglegeneration sequencing in infectious diseases. BMC Infect Dis 2021, 21 (1), 62. DOI: 10.1186/s12879-020-05746-5 From NLM Medline.

(7) Pipoli da Fonseca, J.; Kornobis, E.; Turc, E.; Enouf, V.; Lemee, L.; Cokelaer, T.; Monot, M. Capturing SARS-CoV-2 from patient samples with low viral abundance: a comparative analysis. Sci Rep 2022, 12 (1), 19274. DOI: 10.1038/s41598-022-23422-3 From NLM Medline.

(8) Meyer, F.; Fritz, A.; Deng, Z. L.; Koslicki, D.; Lesker, T. R.; Gurevich, A.; Robertson, G.; Alser, M.; Antipov, D.; Beghini, F.; et al. Critical Assessment of Metagenome Interpretation: the second round of challenges. Nat Methods 2022, 19 (4), 429–440. DOI: 10.1038/s41592-022-01431-4 From NLM Medline.

(9) Kubik, S.; Marques, A. C.; Xing, X.; Silvery, J.; Bertelli, C.; De Maio, F.; Pournaras, S.; Burr, T.; Duffourd, Y.; Siemens, H.; et al. Recommendations for accurate genotyping of SARS-CoV-2 using amplicon-based sequencing of clinical samples. Clin Microbiol Infect 2021, 27 (7), 1036 e1031–1036 e1038. DOI: 10.1016/j.cmi.2021.03.029 From NLM Medline.

(10) Brinkmann, A.; Uddin, S.; Ulm, S. L.; Pape, K.; Forster, S.; Enan, K.; Nourlil, J.; Krause, E.; Schaade, L.; Michel, J.; Nitsche, A. RespiCoV: Simultaneous identification of Severe Acute Respiratory Syndrome Coronavirus 2 (SARS-CoV-2) and 46 respiratory tract viruses and bacteria by amplicon-based Oxford-Nanopore MinION sequencing. PLoS One 2022, 17 (3), e0264855. DOI: 10.1371/journal.pone.0264855 From NLM Medline.

(11) Nagy-Szakal, D.; Couto-Rodriguez, M.; Wells, H. L.; Barrows, J. E.; Debieu, M.; Butcher, K.; Chen, S.; Berki, A.; Hager, C.; Boorstein, R. J.; et al. Targeted Hybridization Capture of SARS-CoV-2 and Metagenomics Enables Genetic Variant Discovery and Nasal Microbiome Insights. Microbiol Spectr 2021, 9 (2), e0019721. DOI: 10.1128/Spectrum.00197-21 From NLM Medline.

(12) Briese, T.; Kapoor, A.; Mishra, N.; Jain, K.; Kumar, A.; Jabado, O. J.; Lipkin, W. I. Virome Capture Sequencing Enables Sensitive Viral Diagnosis and Comprehensive Virome Analysis. mBio 2015, 6 (5), e01491–01415. DOI: 10.1128/mBio.01491-15 From NLM Medline.

(13) Chen, C.; Li, J.; Di, L.; Jing, Q.; Du, P.; Song, C.; Li, J.; Li, Q.; Cao, Y.; Xie, X. S.; et al. MINERVA: A Facile Strategy for SARS-CoV-2 Whole-Genome Deep Sequencing of Clinical Samples. Mol Cell 2020, 80 (6), 1123–1134 e1124. DOI: 10.1016/j.molcel.2020.11.030 From NLM Medline.

(14) Cardozo, K. H. M.; Lebkuchen, A.; Okai, G. G.; Schuch, R. A.; Viana, L. G.; Olive, A. N.; Lazari, C. D. S.; Fraga, A. M.; Granato, C. F. H.; Pintao, M. C. T.; Carvalho, V. M. Establishing a mass spectrometry-based system for rapid detection of SARS-CoV-2 in large clinical sample cohorts. Nat Commun 2020, 11 (1), 6201. DOI: 10.1038/s41467-020-19925-0 From NLM Medline.

(15) Renuse, S.; Vanderboom, P. M.; Maus, A. D.; Kemp, J. V.; Gurtner, K. M.; Madugundu, A. K.; Chavan, S.; Peterson, J. A.; Madden, B. J.; Mangalaparthi, K. K.; et al. A mass spectrometry-based targeted assay for detection of SARS-CoV-2 antigen from clinical specimens. EBioMedicine 2021, 69, 103465. DOI: 10.1016/j.ebiom.2021.103465 From NLM Medline.

(16) Van Puyvelde, B.; Van Uytfanghe, K.; Tytgat, O.; Van Oudenhove, L.; Gabriels, R.; Bouwmeester, R.; Daled, S.; Van Den Bossche, T.; Ramasamy, P.; Verhelst, S.; et al. Cov-MS: A Community-Based Template Assay for Mass-Spectrometry-Based Protein Detection in SARS-CoV-2 Patients. JACS Au 2021, 1 (6), 750–765. DOI: 10.1021/jacsau.1c00048 From NLM PubMed-not-MEDLINE.

(17) Grossegesse, M.; Hartkopf, F.; Nitsche, A.; Schaade, L.; Doellinger, J.; Muth, T. Perspective on Proteomics for Virus Detection in Clinical Samples. J Proteome Res 2020, 19 (11), 4380–4388. DOI: 10.1021/acs.jproteome.0c00674 From NLM Medline.

(18) Zecha, J.; Lee, C. Y.; Bayer, F. P.; Meng, C.; Grass, V.; Zerweck, J.; Schnatbaum, K.; Michler, T.; Pichlmair, A.; Ludwig, C.; Kuster, B. Data, Reagents, Assays and Merits of Proteomics for SARS-CoV-2 Research and Testing. Mol Cell Proteomics 2020, 19 (9), 1503–1522. DOI: 10.1074/mcp.RA120.002164 From NLM Medline.

(19) Messner, C. B.; Demichev, V.; Bloomfield, N.; Yu, J. S. L.; White, M.; Kreidl, M.; Egger, A. S.; Freiwald, A.; Ivosev, G.; Wasim, F.; et al. Ultra-fast proteomics with Scanning SWATH. Nat Biotechnol 2021, 39 (7), 846–854. DOI: 10.1038/s41587-021-00860-4 From NLM Medline.

(20) Manon, B.; Isabelle, F. G.; Ingrid, A. I. V.-V.; Merel, F. A. S.; Rob, S.; Esther, H.; Rene, B.; Christoph, S.; Hans, C.; Theo, M. L.; et al. Proteome2virus: Shotgun mass spectrometry data analysis pipeline for virus identification. Journal of Clinical Virology Plus 2023, 3 (2), 100147. DOI: 10.1016/j.jcvp.2023.100147.

(21) Puyskens, A.; Krause, E.; Michel, J.; Nubling, C. M.; Scheiblauer, H.; Bourquain, D.; Grossegesse, M.; Valusenko, R.; Corman, V. M.; Drosten, C.; et al. Establishment of a specimen panel for the decentralised technical evaluation of the sensitivity of 31 rapid diagnostic tests for SARS-CoV-2 antigen, Germany, September 2020 to April 2021. Euro Surveill 2021, 26 (44). DOI: 10.2807/1560-7917.ES.2021.26.44.2100442 From NLM Medline.

(22) Zougman, A.; Selby, P. J.; Banks, R. E. Suspension trapping (STrap) sample preparation method for bottom-up proteomics analysis. Proteomics 2014, 14 (9), 1006–1000. DOI: 10.1002/pmic.201300553 From NLM Medline.

(23) Skowronek, P.; Thielert, M.; Voytik, E.; Tanzer, M. C.; Hansen, F. M.; Willems, S.; Karayel, O.; Brunner, A. D.; Meier, F.; Mann, M. Rapid and In-Depth Coverage of the (Phospho-)Proteome With Deep Libraries and Optimal Window Design for dia-PASEF. Mol Cell Proteomics 2022, 21 (9), 100279. DOI: 10.1016/j.mcpro.2022.100279 From NLM Medline.

(24) UniProt, C. UniProt: the Universal Protein Knowledgebase in 2023. Nucleic Acids Res 2023, 51 (D1), D523–D531. DOI: 10.1093/nar/gkac1052 From NLM Medline.

(25) Demichev, V.; Messner, C. B.; Vernardis, S. I.; Lilley, K. S.; Ralser, M. DIA-NN: neural networks and interference correction enable deep proteome coverage in high throughput. Nat Methods 2020, 17 (1), 41–44. DOI: 10.1038/s41592-019-0638-x From NLM Medline.

(26) Silva, J. C.; Gorenstein, M. V.; Li, G. Z.; Vissers, J. P.; Geromanos, S. J. Absolute quantification of proteins by LCMSE: a virtue of parallel MS acquisition. Mol Cell Proteomics 2006, 5 (1), 144–156. DOI: 10.1074/mcp.M500230-MCP200 From NLM Medline.

(27) Grossegesse, M.; Leupold, P.; Doellinger, J.; Schaade, L.; Nitsche, A. Inactivation of Coronaviruses during Sample Preparation for Proteomics Experiments. J Proteome Res 2021, 20 (9), 4598–4602. DOI: 10.1021/acs.jproteome.1c00320 From NLM Medline.

(28) Mun, D. G.; Vanderboom, P. M.; Madugundu, A. K.; Garapati, K.; Chavan, S.; Peterson, J. A.; Saraswat, M.; Pandey, A. DIA-Based Proteome Profiling of Nasopharyngeal Swabs from COVID-19 Patients. J Proteome Res 2021, 20 (8), 4165–4175. DOI: 10.1021/acs.jproteome.1c00506 From NLM Medline.

(29) Viode, A.; Smolen, K. K.; Fatou, B.; Wurie, Z.; Van Zalm, P.; Konde, M. K.; Keita, B. M.; Ablam, R. A.; Fish, E. N.; Steen, H. Plasma Proteomic Analysis Distinguishes Severity Outcomes of Human Ebola Virus Disease. mBio 2022, 13 (3), e0056722. DOI: 10.1128/mbio.00567-22 From NLM Medline.

(30) Warinner, C.; Korzow Richter, K.; Collins, M. J. Paleoproteomics. Chem Rev 2022, 122 (16), 13401–13446. DOI: 10.1021/acs.chemrev.1c00703 From NLM Medline.

(31) Castaneda-Mogollon, D.; Kamaliddin, C.; Oberding, L.; Liu, Y.; Mohon, A. N.; Faridi, R. M.; Khan, F.; Pillai, D. R. A metagenomics workflow for SARS-CoV-2 identification, co-pathogen detection, and overall diversity. J Clin Virol 2021, 145, 105025. DOI: 10.1016/j.jcv.2021.105025 From NLM Medline.

(32) Gauthier, N. P. G.; Nelson, C.; Bonsall, M. B.; Locher, K.; Charles, M.; MacDonald, C.; Krajden, M.; Chorlton, S. D.; Manges, A. R. Nanopore metagenomic sequencing for detection and characterization of SARS-CoV-2 in clinical samples. PLoS One 2021, 16 (11), e0259712. DOI: 10.1371/journal.pone.0259712 From NLM Medline.

(33) Mostafa, H. H.; Fissel, J. A.; Fanelli, B.; Bergman, Y.; Gniazdowski, V.; Dadlani, M.; Carroll, K. C.; Colwell, R. R.; Simner, P. J. Metagenomic Next-Generation Sequencing of Nasopharyngeal Specimens Collected from Confirmed and Suspect COVID-19 Patients. mBio 2020, 11 (6). DOI: 10.1128/mBio.01969-20 From NLM Medline.

(34) Babiker, A.; Bradley, H. L.; Stittleburg, V. D.; Ingersoll, J. M.; Key, A.; Kraft, C. S.; Waggoner, J. J.; Piantadosi, A. Metagenomic Sequencing To Detect Respiratory Viruses in Persons under Investigation for COVID-19. J Clin Microbiol 2020, 59 (1). DOI: 10.1128/JCM.02142-20 From NLM Medline.

(35) Jurasz, H.; Pawlowski, T.; Perlejewski, K. Contamination Issue in Viral Metagenomics: Problems, Solutions, and Clinical Perspectives. Front Microbiol 2021, 12, 745076. DOI: 10.3389/fmicb.2021.745076 From NLM PubMed-not-MEDLINE.

(36) Stewart, H. I.; Grinfeld, D.; Giannakopulos, A.; Petzoldt, J.; Shanley, T.; Garland, M.; Denisov, E.; Peterson, A. C.; Damoc, E.; Zeller, M.; et al. Parallelized Acquisition of Orbitrap and Astral Analyzers Enables High-Throughput Quantitative Analysis. Anal Chem 2023, 95 (42), 15656–15664. DOI: 10.1021/acs.analchem.3c02856 From NLM PubMed-not-MEDLINE.

(37) Guzman, U. H.; Martinez-Val, A.; Ye, Z.; Damoc, E.; Arrey, T. N.; Pashkova, A.; Renuse, S.; Denisov, E.; Petzoldt, J.; Peterson, A. C.; et al. Ultra-fast label-free quantification and comprehensive proteome coverage with narrow-window data-independent acquisition. Nat Biotechnol 2024. DOI: 10.1038/s41587-023-02099-7 From NLM Publisher.

(38) van Kampen, J. J. A.; van de Vijver, D.; Fraaij, P. L. A.; Haagmans, B. L.; Lamers, M. M.; Okba, N.; van den Akker, J. P. C.; Endeman, H.; Gommers, D.; Cornelissen, J. J.; et al. Duration and key determinants of infectious virus shedding in hospitalized patients with coronavirus disease-2019 (COVID-19). Nat Commun 2021, 12 (1), 267. DOI: 10.1038/s41467-020-20568-4 From NLM Medline.

(39) Michel, J.; Neumann, M.; Krause, E.; Rinner, T.; Muzeniek, T.; Grossegesse, M.; Hille, G.; Schwarz, F.; Puyskens, A.; Forster, S.; et al. Resource-efficient internally controlled in-house real-time PCR detection of SARS-CoV-2. Virol J 2021, 18 (1), 110. DOI: 10.1186/s12985-021-01559-3 From NLM Medline.

(40) Fomsgaard, A. S.; Rasmussen, M.; Spiess, K.; Fomsgaard, A.; Belsham, G. J.; Fonager, J. Improvements in metagenomic virus detection by simple pretreatment methods. J Clin Virol Plus 2022, 2 (4), 100120. DOI: 10.1016/j.jcvp.2022.100120 From NLM PubMed-not-MEDLINE.

(41) Blume, J. E.; Manning, W. C.; Troiano, G.; Hornburg, D.; Figa, M.; Hesterberg, L.; Platt, T. L.; Zhao, X.; Cuaresma, R. A.; Everley, P. A.; et al. Rapid, deep and precise profiling of the plasma proteome with multi-nanoparticle protein corona. Nat Commun 2020, 11 (1), 3662. DOI: 10.1038/s41467-020-17033-7 From NLM Medline.

(42) Viode, A.; van Zalm, P.; Smolen, K. K.; Fatou, B.; Stevenson, D.; Jha, M.; Levy, O.; Steen, J.; Steen, H.; Network, I. A simple, time- and cost-effective, high-throughput depletion strategy for deep plasma proteomics. Sci Adv 2023, 9 (13), eadf9717. DOI: 10.1126/sciadv.adf9717 From NLM Medline.

